## Supplementary materials for "Preventing iatrogenic HCV infection: A quantitative risk assessment based on observational data in an Egyptian hospital"

**Supplementary appendix**

***Table S1****:* Characteristics of each group found after sequence analysis. …..…………………………………..……1

***Figure S1:*** Procedure–specific risks …………………………..……………..……...…………...……...…………3

***Figure S2:*** Point Biserial Correlation………………………………………………………………………………3

***Text S1*:** Mode calculation ………………………………………………………………………………………....4

***Table S2****:* Patient and ward-focused strategies ………………..………………………………………………...….4

***Table S3****:* Sensitivity analysis for the logistic regression …………………..……………..….…….…...…………5

**Table S1.** Characteristics of each group found after sequence analysis.

| *Characteristic* | *Group 1, N = 356* | *Group 2, N = 54* | *Group 3, N = 14* | *Group 4, N = 76* | *p-value* | *Overall, N = 500* |
| --- | --- | --- | --- | --- | --- | --- |
| ***Gender*** |  |  |  |  | *0·6* |  |
| *Female* | *151 (42%)* | *19 (35%)* | *6 (43%)* | *36 (47%)* |  | *212 (42%)* |
| *Male* | *205 (58%)* | *35 (65%)* | *8 (57%)* | *40 (53%)* |  | *288 (58%)* |
| ***Age*** | *43 (30, 58)* | *42 (29, 57)* | *64 (46, 67)* | *53 (37, 64)* | ***0·006*** | *45 (30, 60)* |
| ***Education level*** |  |  |  |  | *0·058* |  |
| *No formal education* | *156 (44%)* | *16 (30%)* | *8 (57%)* | *44 (58%)* |  | *224 (45%)* |
| *Primary or preparatory school* | *53 (15%)* | *10 (19%)* | *2 (14%)* | *11 (14%)* |  | *76 (15%)* |
| *Secondary school or higher* | *147 (41%)* | *28 (52%)* | *4 (29%)* | *21 (28%)* |  | *200 (40%)* |
| ***Marital status*** |  |  |  |  | *0·6* |  |
| *Single* | *54 (15%)* | *8 (15%)* | *2 (14%)* | *7 (9·2%)* |  | *71 (14%)* |
| *Maried* | *268 (75%)* | *38 (70%)* | *10 (71%)* | *58 (76%)* |  | *374 (75%)* |
| *Widow* | *27 (7·6%)* | *5 (9·3%)* | *2 (14%)* | *7 (9·2%)* |  | *41 (8·2%)* |
| *Divorced* | *7 (2·0%)* | *3 (5·6%)* | *0 (0%)* | *4 (5·3%)* |  | *14 (2·8%)* |
| ***Localization*** |  |  |  |  | *0·12* |  |
| *Cairo* | *244 (69%)* | *37 (69%)* | *8 (57%)* | *42 (55%)* |  | *331 (66%)* |
| *Other governate* | *110 (31%)* | *17 (31%)* | *6 (43%)* | *34 (45%)* |  | *167 (34%)* |
| *Unknown* | *2* | *0* | *0* | *0* |  | *2* |
| ***Patient Hospitalized before*** | *258 (72%)* | *40 (74%)* | *12 (86%)* | *66 (88%)* | ***0·031*** | *376 (75%)* |
| *Unknown* | *0* | *0* | *0* | *1* |  | *1* |
| ***Source of admission*** |  |  |  |  | *0·082* |  |
| *Outpatient clinic* | *151 (42%)* | *32 (59%)* | *5 (36%)* | *38 (50%)* |  | *226 (45%)* |
| *Emergency room* | *205 (58%)* | *22 (41%)* | *9 (64%)* | *38 (50%)* |  | *274 (55%)* |
| ***Hospital at recruitment*** |  |  |  |  | ***<0·001*** |  |
| *Surgery hospital* | *219 (62%)* | *54 (100%)* | *1 (7·1%)* | *0 (0%)* |  | *274 (55%)* |
| *Internal medicine hospital* | *137 (38%)* | *0 (0%)* | *13 (93%)* | *76 (100%)* |  | *226 (45%)* |
| ***Status at the end of following*** |  |  |  |  | ***<0·001*** |  |
| *Deceased* | *0 (0%)* | *0 (0%)* | *5 (36%)* | *0 (0%)* |  | *5 (1·0%)* |
| *Discharged* | *356 (100%)* | *54 (100%)* | *9 (64%)* | *76 (100%)* |  | *495 (99%)* |
| ***Average estimated risk of HVC infection (%)*** | *0·04% (0·02%, 0·07%)* | *0·07% (0·05%, 0·11%)* | *0·47% (0·08%, 0·82%)* | *0·12% (0·03%, 0·30%)* | ***<0·001*** | *0·04% (0·03%, 0·09%)* |
| ***Duration of hospitalization (days)*** | *2·3 (1·7, 3·3)* | *6·4 (5·4, 9·0)* | *20·4 (17·5, 23·2)* | *8·7 (7·2, 10·9)* | ***<0·001*** | *3·1 (1·9, 5·8)* |
| ***Number of procedures*** | *6 (4, 8)* | *14 (9, 23)* | *43 (15, 77)* | *12 (5, 23)* | ***<0·001*** | *7 (4, 11)* |
| **Reason for hospitalisation** |  |  |  |  | ***<0·001*** |  |
| General surgery | *152 (43%)* | *26 (48%)* | *2 (14%)* | *1 (1·3%)* |  | *181 (36%)* |
| Special surgery | *54 (15%)* | *23 (43%)* | *2 (14%)* | *22 (29%)* |  | *101 (20%)* |
| General IM | *30 (8·4%)* | *0 (0%)* | *2 (14%)* | *12 (16%)* |  | *44 (8·8%)* |
| Special IM | *78 (22%)* | *4 (7·4%)* | *6 (43%)* | *33 (43%)* |  | *121 (24%)* |
| Liver/ GIT complaint | *42 (12%)* | *1 (1·9%)* | *2 (14%)* | *8 (11%)* |  | *53 (11%)* |
| *n (%); Median (IQR)* | | | | | | |
| 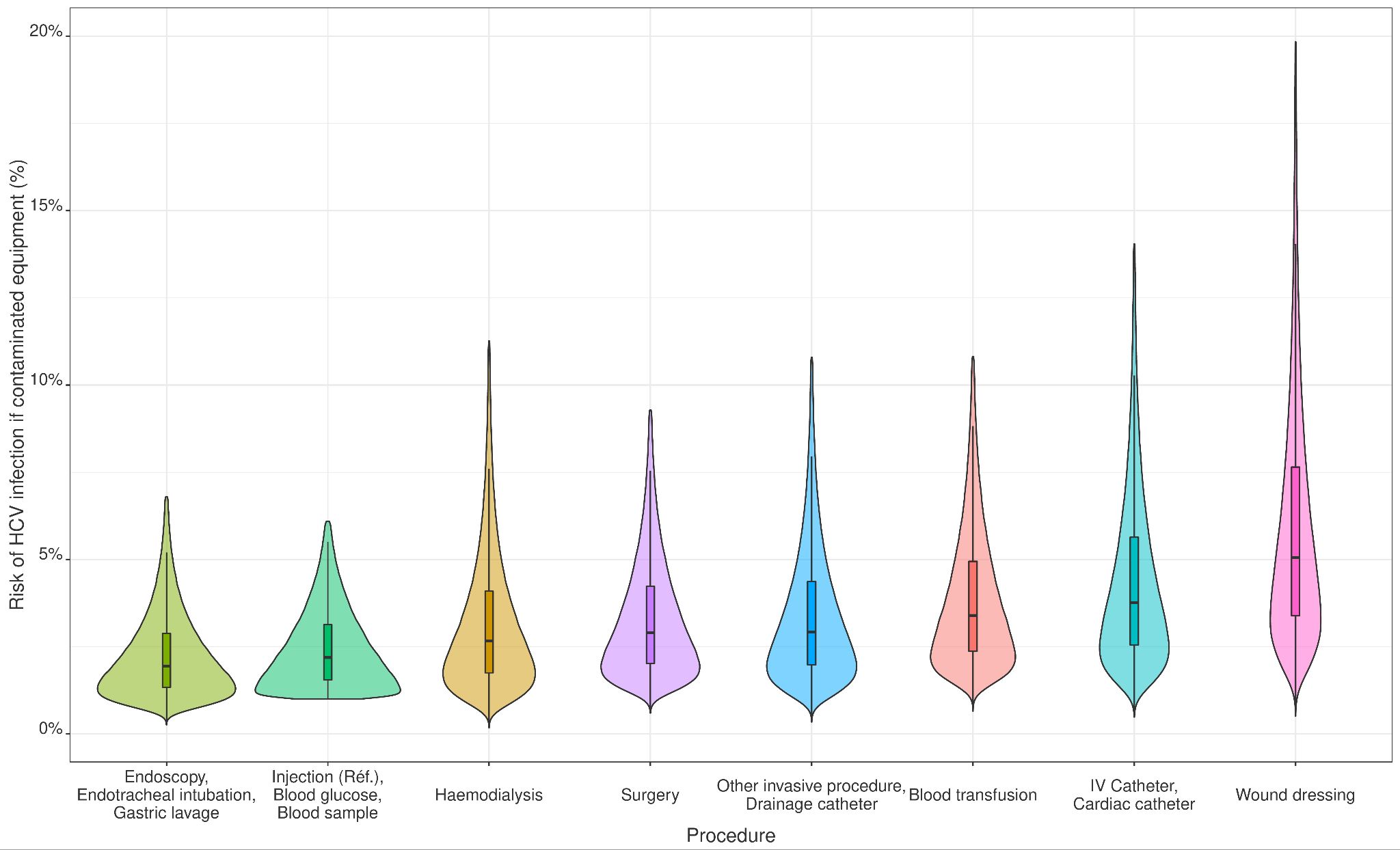  **Figure S1.** Distributions of the procedure–specific risks of HCV infection in case of contaminated equipment  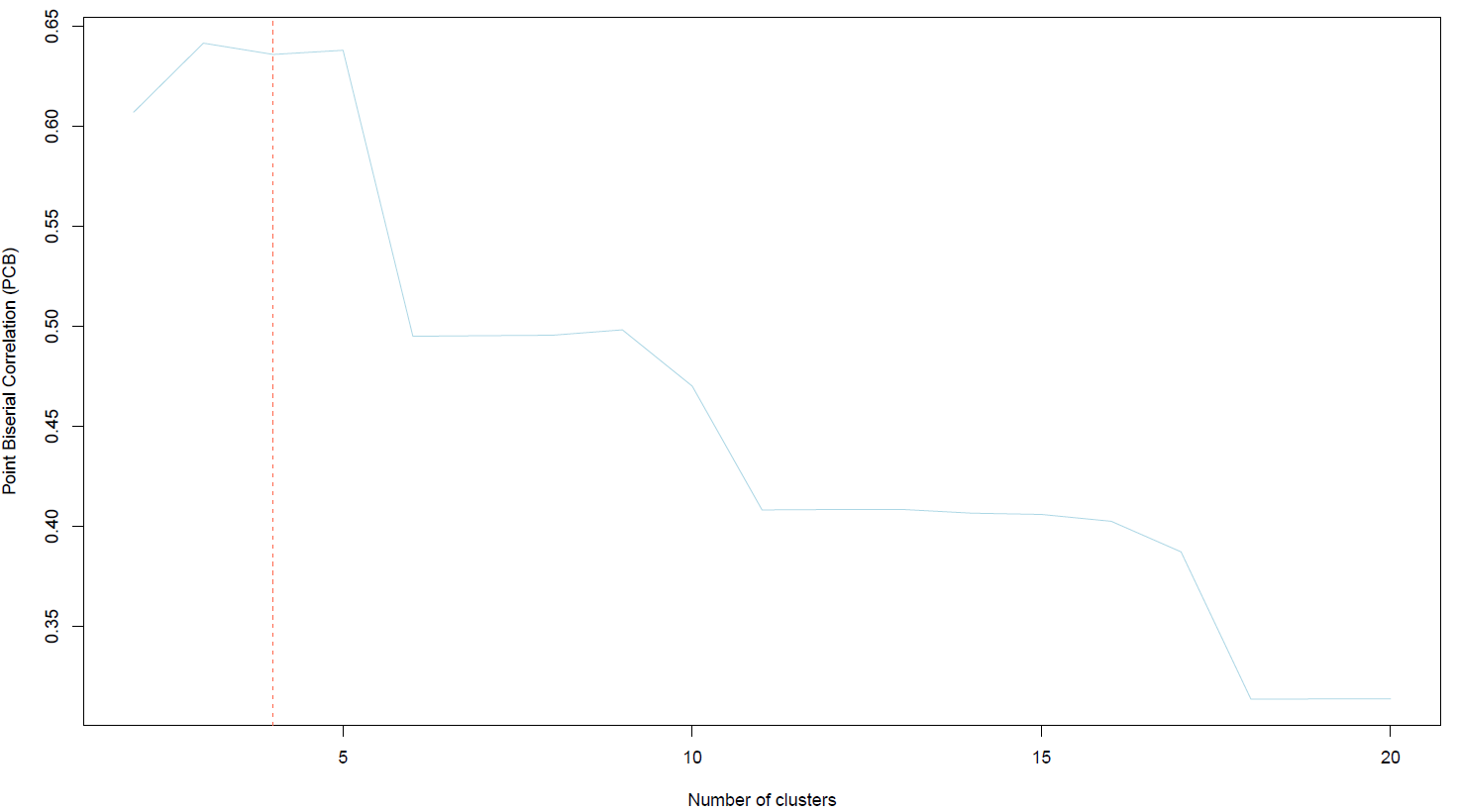 | | | | | | |

**Figure S2.** Point Biserial Correlation (PBC) for 1 to 20 clusters. PBC was very similar for 3, 4 and 5 partitions. Therefore, we chose to build 4 clusters of patients (vertical dashed line).

**Text S1.** Mode calculation

The median of a PERT distribution can be approximated by $\frac{a+6b+c}{8}$ where $a$ and $c$ correspond to the minimum and maximum values of the risk and $b$ to the mode of the distribution. Knowing that $a = 1$ and $c = 9$·$2$, and assuming that the median is equal to 2·2%, we need to solve $\frac{a+6b+c}{8}=2\cdot2$ which gives $b = 1\cdot23$ .

**Table S2.** Summary of the impact of patient and ward-focused strategies on the risk of HCV infection during hospitalization.

| **Intervention** | **Average risk reduction**  **(Std. error)** | **Average risk**  **(Std. error)** | **Number of patients concerned by intervention (% of total)** | **Wards concerned for the ward-focused scenario (Number)** |
| --- | --- | --- | --- | --- |
| *Patient-focused*  *(Model-based selection)* | 80·12%  (9·14x10^-4^) | 0·023%  (8·46x10^-5^) | 200 (40%) | - |
|  | 79·09%  (4·95x10^-4^) | 0·025%  (2·91x10^-5^) | 175 (35%) | - |
|  | 73·01%  (9·00x10^-4^) | 0·031%  (2·71x10^-4^) | 150 (30%) | - |
|  | 56·71%  (2·08x10^-3^) | 0·049%  (1·78x10^-4^) | 125 (25%) | - |
|  | 52·51%  (1·22x10^-3^) | 0·058%  (1·86x10^-4^) | 100 (20%) | - |
| *Patient-focused*  *(Random selection)* | 63·39%  (9·29x10^-3^) | 0·042%  (1·03x10^-3^) | 200 (40%) | - |
|  | 55·10%  (1·07x10^-2^) | 0·052%  (1·20x10^-3^) | 175 (35%) | - |
|  | 49·56%  (1·31x10^-3^) | 0·058%  (1·29x10^-3^) | 150 (30%) | - |
|  | 42·77%  (1·27x10^-2^) | 0·066%  (1·20x10^-3^) | 125 (25%) | - |
|  | 34·95%  (1·45x10^-2^) | 0·075%  (1·75x10^-3^) | 100 (20%) | - |
| *Ward-focused* | 76·93%  (4·48x10^-4^) | 0·027%  (8·26x10^-5^) | 203 (40.6%) | Rheumatology, Tropical medicine, ER ICU, GIT and endoscopy, Cardiology,Neurology,Immunology,Urosurgery,Ophtalmology (9) |
|  | 72·06%  (7·41x10^-4^) | 0·032%  (7·23x10^-5^) | 182 (36.4%) | Rheumatology, Tropical medicine, ER ICU, GIT and endoscopy, Cardiology,Neurology,Immunology (7) |
|  | 62·97%  (7·47x10^-4^) | 0·043%  (7·83x10^-5^) | 159 (31.8%) | Rheumatology, Tropical medicine, ER ICU, GIT and endoscopy, Cardiology (5) |
|  | 56·02%  (1·07x10^-3^) | 0·051%  (1·21x10^-4^) | 82(16.4%) | Rheumatology, Tropical medicine, ER ICU, GIT and endoscopy (4) |

**Table S3.** Sensitivity analysis for the cut-off value of the risk considered in the logistic regression.

| **Quantile proba. for overall risk in baseline scenario** | **Cut-off value for the risk (%)** | **AUC [CI95%]** | **Specificity [CI95%]** | **Sensitivity [CI95%]** | **Precision [CI95%]** | **Informedness** |
| --- | --- | --- | --- | --- | --- | --- |
| 0·60 | 0·056% | *0·76 [0·69-0·83]* | 0·62[0·54-0·71] | 0·69[0·58-0·78] | 0·55[0·45-0·64] | 0·31 |
| 0·65 | 0·066% | *0·80[0·73-0·87]* | 0·76[0·68-0·83] | 0·70[0·58-0·79] | 0·61[0·5-0·71] | 0·46 |
| 0·70 | 0·076% | *0·82[0·75-0·89]* | 0·72[0·64-0·79] | 0·73[0·61-0·83] | 0·53[0·42-0·63] | 0·45 |
| 0·75 | 0·096% | *0·79[0·71-0·87]* | 0·73[0·65-0·79] | 0·68[0·54-0·79] | 0·45[0·35-0·57] | 0·41 |
| 0·80 | 0·12% | *0·78[0·69-0·87]* | 0·71[0·64-0·78] | 0·70[0·55-0·82] | 0·38[0·28-0·49] | 0·41 |
| 0·85 | 0·16% | *0·77[0·67-0·87]* | 0·75[-0·68-0·81] | 0·67[0·39-0·73] | 0·29[0·19-0·41] | 0·32 |
| 0·90 | 0·25% | *0·82[0·7-0·94]* | 0·64[0·57-0·71] | 0·90[0·7-0·97] | 0·22[0·14-0·32] | 0·54 |
| 0·95 | 0·49% | *0·72[0·54-0·9]* | 0·75[0·68-0·8] | 0·60[0·31-0·83] | 0·11[0·05-0·22] | 0·35 |
